# A systematic review of Nature-based interventions for neurological disorders

**DOI:** 10.1101/2025.06.21.25330058

**Authors:** Christian Kovaleski, Najma Ahmed, Cat Huckle, James Bashford

**Author notes:** Correspondence to: Dr James Bashford, Maurice Wohl Clinical Neuroscience Institute, 5 Cutcombe Rd, London, SE5 9RT, UK., Website: https://www.healthconservation.org Twitter: @healthconserve.

## Abstract

The human nervous system evolved to optimise its interactions with the natural world. However, as civilisations over recent millennia have developed, the human brain has become steadily disconnected from these natural stimuli. Consequently, detrimental effects on individual health are increasingly recognised. Strategies that promote reconnection with Nature have yielded significant gains when it comes to psychological and general physical health, although much less is known about their direct impact on neurological health. In this systematic review, we identified 17 studies that assessed at least one Nature-based intervention to treat any neurological disease. The predominant study design was quasi-experimental (n=9). Interventions included horticultural therapy, petal arranging and farm visits. The majority of studies had a moderate risk of bias. While 88% (n=15) of studies reported improvements in measures such as quality of life, mood, agitation, apathy or cognitive function, there was such an overrepresentation of studies recruiting patients with dementia (94%; n=16) that for many common neurological disorders, including epilepsy, migraine and multiple sclerosis, the current literature leaves us none the wiser. Considering the extensive crosstalk that exists between physical and psychological disease, we argue that the potential benefit of formal ‘Nature Prescriptions’ as an accessible, cheap and harmless endorsement of Mother Nature’s healing hand warrants greater attention in neurology. Indeed, this aligns with the wider societal and environmental need for humans to reconnect with the natural world.

## INTRODUCTION

In this age of modern medicine, it is increasingly clear that tackling ill health requires more than understanding the associated pathogens or genetic risk factors (<u>Stigsdotter et al., 2011</u>). There is growing appreciation for the detrimental impact of certain aspects of modern life, such as chronic stress, with their connection to physical and psychological diseases, and how these can be detrimental to quality of life (<u>Oh et al., 2020</u>). With this in mind, a holistic approach is vital when attempting to improve public health. One valuable, yet underrated, aspect is the therapeutic benefit of Nature. As such, Nature-based interventions, Nature Prescriptions and Green Prescriptions have gained increased attention as a means to harness the healing benefits of the natural environment (<u>Twohig-Bennett and Jones, 2018</u>) (<u>Adewuyi et al., 2023</u>). In this systematic review, we identified the studies employing any form of Nature-based intervention for the management of neurological disorders.

Neurological disorders are the leading cause of disability and the second leading cause of death worldwide, with this burden expected to rise across the next few decades (<u>Feigin et al., 2020</u>). These conditions are strongly linked to mental ill health, with reduced health-related quality of life being driven by high rates of depression and anxiety, especially in disorders like epilepsy, where psychiatric comorbidities affect up to 50% of patients (<u>Prisnie et al., 2018</u>, <u>Salpekar and Mula, 2019</u>). Despite this, many patients lack access to appropriate psychological support, highlighting the need for novel interventions that address such comorbidities (<u>Gandy, 2023</u>). Thus, we are interested in discovering whether exposure to Nature could be an effective means to address this overarching issue in neurology.

Nature-based interventions refer to ‘programmes, activities or strategies that aim to engage people in Nature–based experiences with the specific goal of achieving improved health and wellbeing’. (<u>Shanahan et al., 2019</u>). These can be broadly categorised into two groups: those that change the environment in which the target demographic lives (such as the provision of greenspaces/gardens in hospitals), and those that aim to change people’s behaviour through interaction with Nature (such as care farms, Nature play) (<u>Shanahan et al., 2019</u>). This deliverance of a ‘dose of Nature’ is thought to generate the positive outcomes reported.

Prior evidence links greater exposure to Nature with improvements in health and social wellbeing outcomes (<u>Shanahan et al., 2019</u>, <u>White et al., 2019</u>). A recent meta-analysis suggests that Nature-based interventions are successful in providing mental health benefits, such as improving positive affect (<u>Coventry et al., 2021</u>). Further evidence also promotes their positive impact on physical health and attitudes towards Nature (<u>Silva et al., 2023</u>). Additionally, Nature-based interventions may provide benefits beyond health. Case studies highlight reduced public service use (<u>Pretty and Barton, 2020</u>) and reduced costs (e.g., fewer drug prescriptions) as some of the potential economic benefits (<u>Bloomfield, 2017</u>). Socially, participants report benefits such as ‘improvements in their social skills’ and an ‘improved sense of individual worth and of agency’ (<u>Bloomfield, 2017</u>). However, whilst these provide good support for the holistic efficacy of Nature-based interventions, the current idea of a ‘dose of Nature’ is likely to be too simplistic to address the dynamic and complex ways that individuals interact with the natural world (<u>Robinson and Breed, 2019</u>).

To quantify the complex relationship between humans and the natural world, a number of empirical scales have been proposed. What these scales share is their ability to measure the subjective connection between individuals and Nature, so-called ‘Nature connectedness’ (<u>Pasca et al., 2017</u>). This is based on E.O. Wilson’s ‘biophilia’ hypothesis, which suggests that an individual’s psychological health is reliant on our species’ innate affinity with Nature (<u>Kellert and Wilson, 1993</u>). In accordance with this theory, there is growing evidence that contact with non-threatening natural environments is associated with a range of positive health outcomes, with the individual’s psychological relationship with Nature being a key part of this process (<u>Martin et al., 2020</u>). Studies also report that those who are more connected to Nature experience more positive affect, vitality and life satisfaction compared to those less connected to Nature (<u>Capaldi et al., 2014</u>). Importantly, wider use of such Nature connectedness scales should facilitate more precise evaluation of Nature-based interventions.

Despite having positive effects on health, the mechanisms that underpin this process are poorly understood. Modern understanding stems from the idea that humans evolved and adapted to function well in the natural environment, and hence these settings provide more intrinsic benefit than post-industrial surroundings. As such, humans instinctively feel more relaxed in these environments, allowing for reduced stress and other positive psycho-physiological responses (<u>Oh et al., 2020</u>). Indeed, the ‘stress reduction theory’ states that exposure to Nature brings about a prompt alleviation of the acute stress response, including normalised blood pressure and reduced feelings of anxiety (<u>Shaffee and Abd Shukor, 2018</u>, <u>Gritzka et al., 2020</u>). In addition, the ‘attention restoration theory’ posits that prolonged periods of attention directed at a narrow set of tasks induce mental fatigue and negative emotional states. Thankfully, these effects can be reversed by natural stimuli, which evoke a sense of ‘quiet fascination’, thereby refreshing attentional levels (<u>Han, 2017</u>, <u>Berto, 2014</u>).

Given these myriad benefits, we set out to identify and assess the outcomes of all interventional studies that have employed at least one kind of Nature-based intervention to treat patients with any neurological disease.

## METHODS

### Study selection and search strategy

This systematic review follows the PRISMA (Preferred Reporting Items for Systematic Review and Meta-Analyses) 2020 Statement (<u>Page et al., 2021</u>). A systematic computerised search was completed by one author (CK) in April 2023 in four databases: Embase (1974–) Ovid MEDLINE(R) ALL (1946–), Global Health (1973 –) APA PsychInfo (1806–). Searches were tailored to the databases and included key text-word terms, phrases and medical subject headings (MESH) terms with “and/or” for “neurologic disease”, “Nature therapy”, “health outcomes” and related. Terms were connected via Boolean operators using “AND” for PEO (Population, Exposure, Outcome) elements, and “OR” for synonymous search terms within each element. A full search strategy is presented as *table A* in the appendix.

Rayyan.ai was used to facilitate the selection process. Records filtered from the initial search were inputted, and possible duplicates were resolved. Titles and abstracts were screened according to our eligibility criteria. Studies included were (a) Primary, interventional studies, where (b) Nature-based interventions were used. Papers not written in English were also excluded. Additionally, qualitative studies and non-empirical papers, such as conference abstracts or systematic reviews were excluded. See *table 1* for full inclusion and exclusion criteria. Full text records remaining were then analysed for their inclusion to the data extraction list. The PRISMA flow chart (*figure 1*) displays the selection process and reasons for exclusions at each stage of screening.

**Figure 1:**
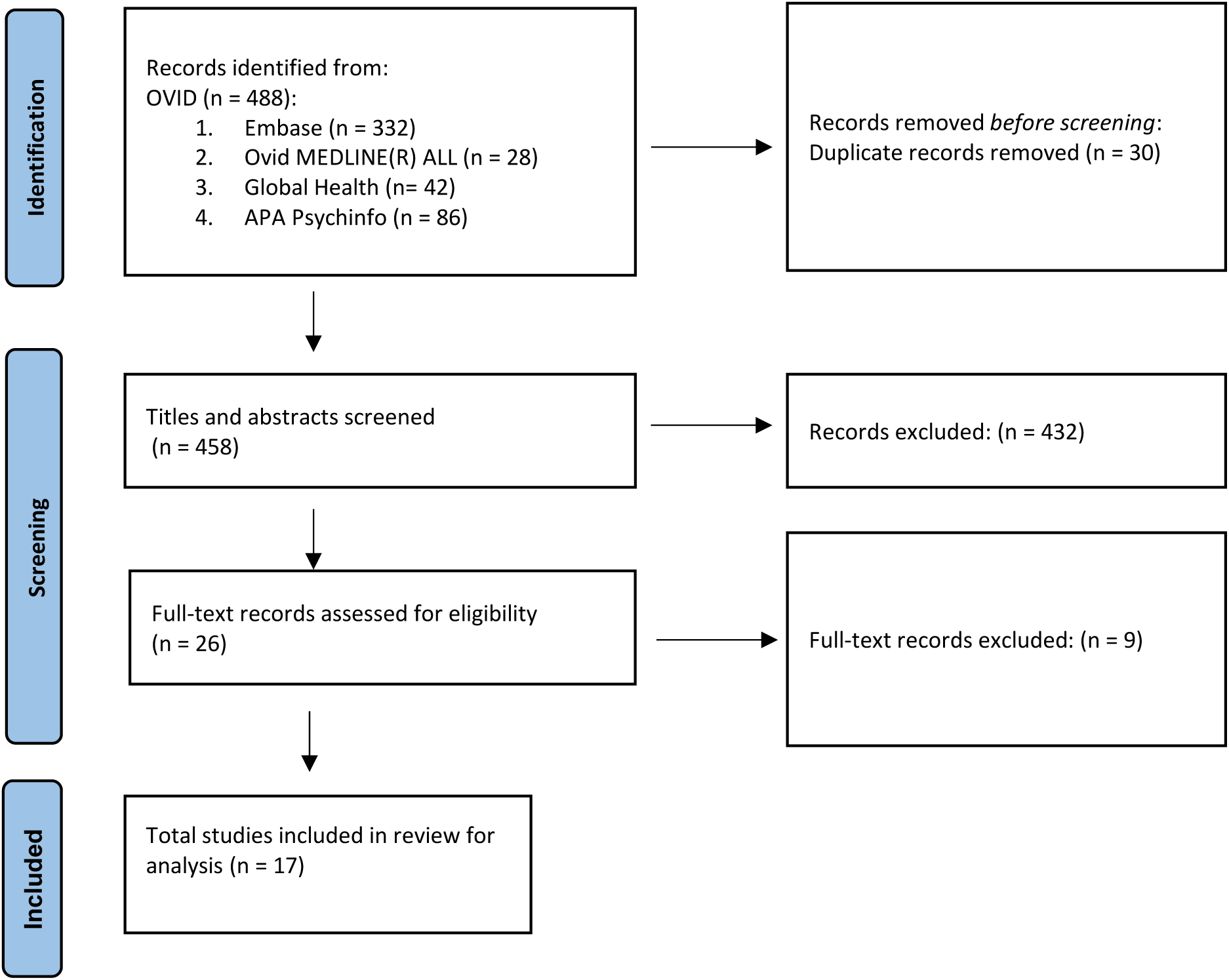
Systematic Review Flow Diagram.

**Table 1.** Eligibility Criteria.

| Inclusion Criteria | Exclusion Criteria |
| --- | --- |
| Primary, interventional studies | Qualitative studies, Non-empirical Studies |
| Nature-based interventions used | Focus of study is not a Nature based intervention |
|  | Papers not written in English. |

We limited this systematic review to peer-reviewed research articles written in English. Duplicates amongst the identified studies were removed. The remaining studies were screened by two authors (CK/NA) for relevance, initially based on title and abstract. Of those remaining, the full-text article was assessed, and any queries were resolved by discussion with the senior author (JB). The final list of selected studies was agreed upon by all authors. The protocol for this review has not previously been published.

### Data extraction and quality assessment

Relevant data were extracted from the selected studies in a table with headings as follows: study details (location, authors), study design (type and number of patients), neurological diseases of interest, Nature-based interventions investigated, outcome measures and findings (positive and negative).

Three tools were used to assess study quality: Newcastle-Ottawa Scale (NOS) (<u>Wells et al., 2000</u>), Adapted NOS for cross-sectional studies (<u>Lippincott Williams & Wilkins, 2019</u>), and Risk of Bias 2 (RoB2) (<u>Sterne et al., 2019</u>). NOS assesses risk of bias in three main domains: selection of the study groups, comparability between groups and assessment of the outcome and exposure. We determined adequate follow-up length to be a period of at least 6 months. The maximum score was 9, and studies with a score of ≥7 were considered to have low risk of bias, those with a score of 4–6 were considered to have moderate bias, and scores of <4 were considered to have high risk of bias. As per recommendations, the scale was evaluated by two independent reviewers (CK/NA) to reduce risk of interpretation bias (<u>Luchini et al., 2017</u>). RoB 2 is structured with five sets of domains of bias, primarily focussing on trial design, conduct and reporting. Judgements for each domain are made following an included algorithm. The domains can be judged as ‘Low risk’, ‘High risk’ or ‘Some concerns’. Overall risk of judgement follows the recommendations set out (<u>Sterne et al., 2019</u>). For all tools, scoring/judgment disagreements were settled by consensus. *Table 2., 3., & 4*. below display the quality assessment results.

**Table 2.** Newcastle Ottawa Scale (NOS)

*Note.* \* = 1 point; \*\* = 2 points
|  | **Selection** |  |  |  | **Comparability** | **Outcome** |  | **Score** |
| --- | --- | --- | --- | --- | --- | --- | --- | --- |
| Author & Date | Representativeness of the sample | Sample size | Non-Response rate | Ascertainment of exposure (max **) | Confounders investigated | Outcome assessment (max **) | Statistical tests | Total |
| 3. | * |  |  | ** | * | * | * | 6/9 |
| 6. | * |  |  | ** | * | * | * | 6/9 |
| 12. | * |  |  | ** |  | * |  | 4/9 |

**Table 3.** Adapted Newcastle Ottawa Scale for Cross-Sectional Studies.

*Note.* \* = 1 point; \*\* = 2 points
|  | **Selection** |  |  |  | **Comparability** | **Outcome** |  |  | **Score** |
| --- | --- | --- | --- | --- | --- | --- | --- | --- | --- |
| Author & Date | Representativeness of exposed cohort | Selection of the non-exposed cohort | Ascertainment of exposure | Demonstration that outcome of interest was not present at start of study | Comparability of cohorts on the basis of the design or analysis (max **) | Outcome assessment (max **) | Follow up length (>6 months) | Adequacy of follow up of cohorts | Total |
| 1. | * |  | * | * |  |  |  | * | 4/9 |
| 2. | * |  | * | * |  |  | * | * | 5/9 |
| 4. | * |  | * | * |  |  | * | * | 5/9 |
| 7. |  | * |  | * | * |  |  |  | 3/9 |
| 11. | * |  | * | * |  |  | * | * | 5/9 |
| 13. | * |  | * | * |  |  |  | * | 4/9 |
| 14. | * | * | * | * | * |  | * | * | 6/9 |
| 15. | * |  | * | * |  |  |  |  | 3/9 |
| 17. | * | * | * | * | * |  |  |  | 5/9 |

**Table 4.**
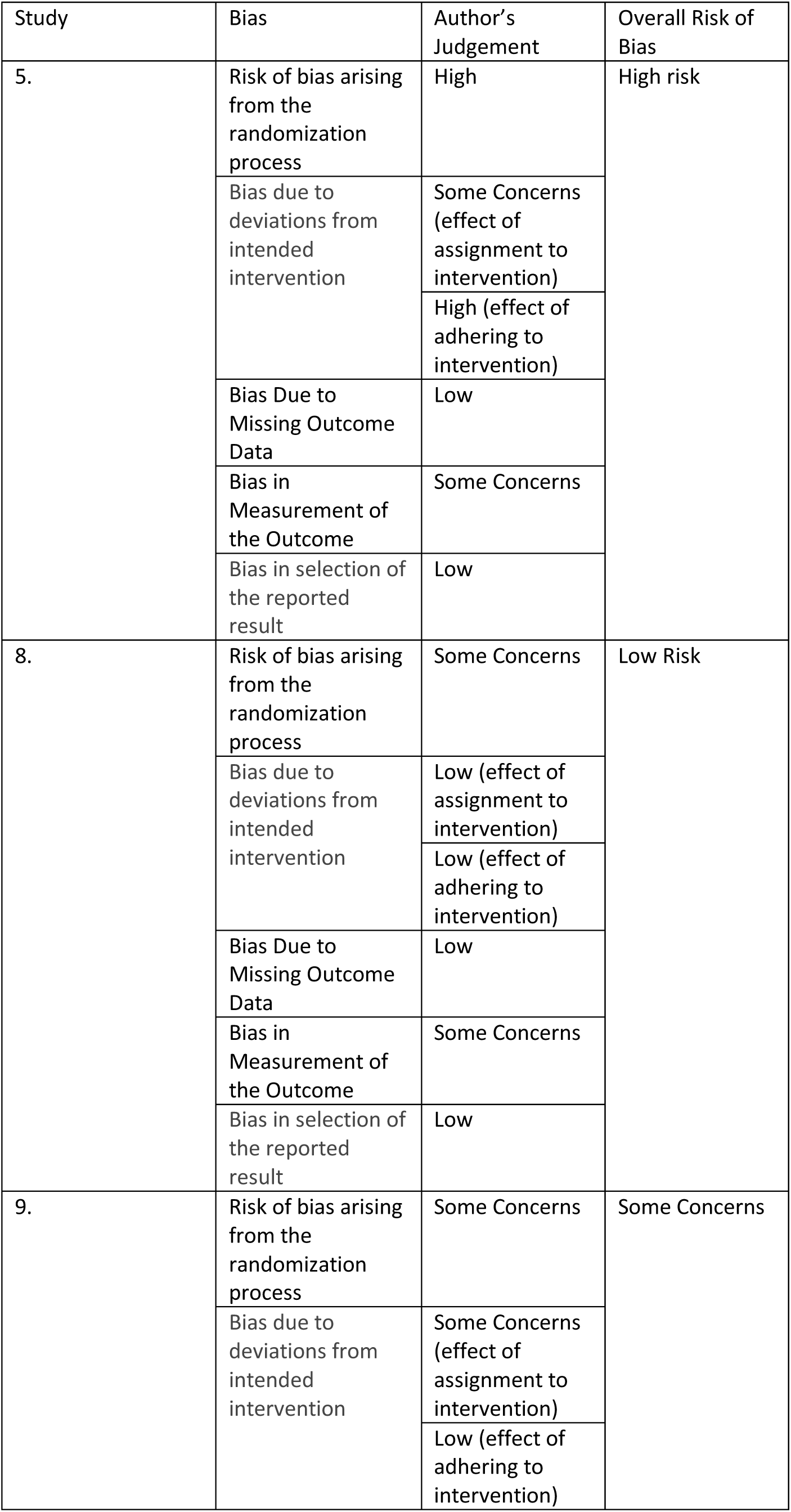
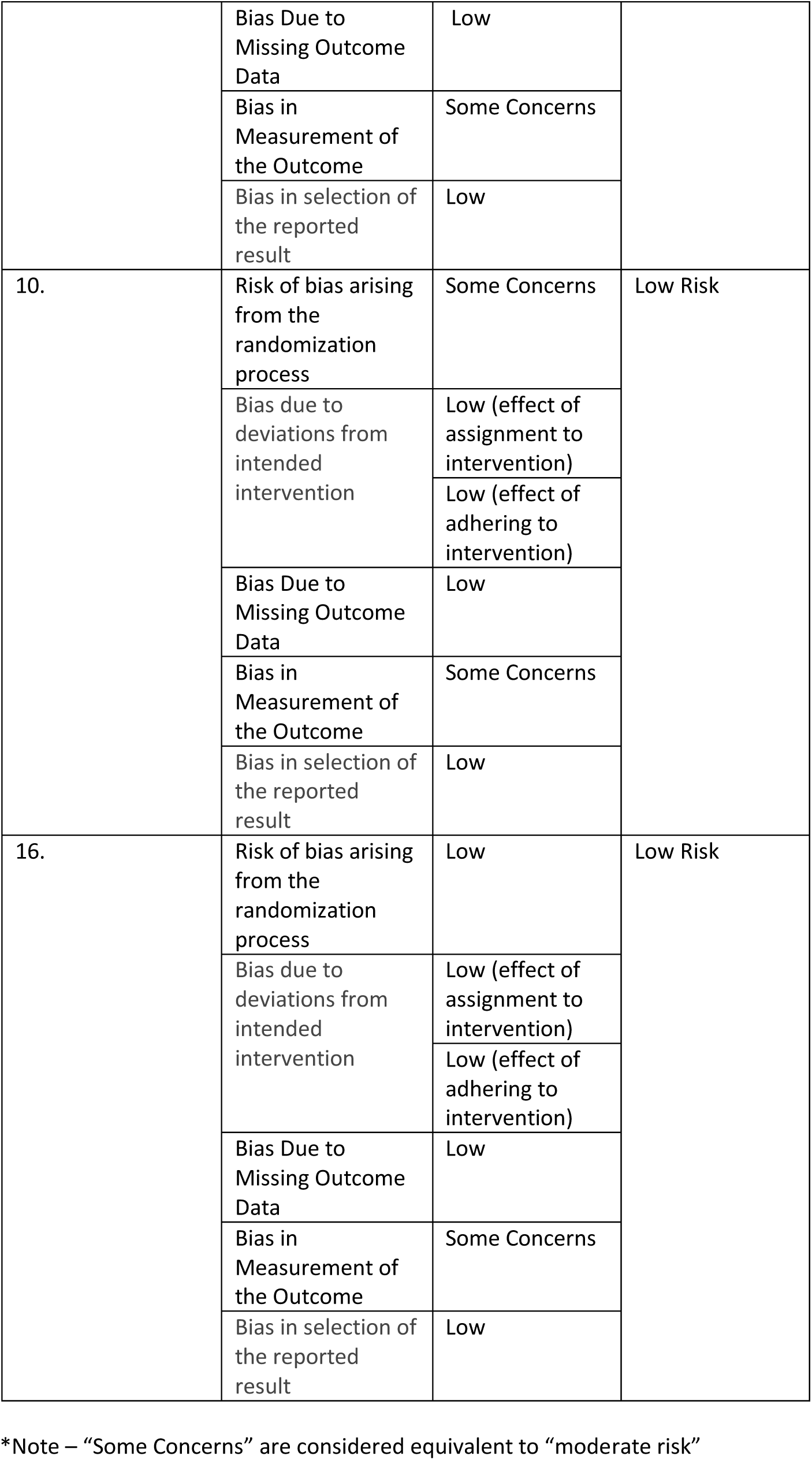
Risk of Bias 2 (RoB2)

| Study | Bias | Author's Judgement | Overall Risk of Bias |
| --- | --- | --- | --- |
| 5. | Risk of bias arising from the randomization process | High | High risk |
|  | Bias due to deviations from intended intervention | Some Concerns (effect of assignment to intervention) |  |
|  |  | High (effect of adhering to intervention) |  |
|  | Bias Due to Missing Outcome Data | Low |  |
|  | Bias in Measurement of the Outcome | Some Concerns |  |
|  | Bias in selection of the reported result | Low |  |
| 8. | Risk of bias arising from the randomization process | Some Concerns | Low Risk |
|  | Bias due to deviations from intended intervention | Low (effect of assignment to intervention) |  |
|  |  | Low (effect of adhering to intervention) |  |
|  | Bias Due to Missing Outcome Data | Low |  |
|  | Bias in Measurement of the Outcome | Some Concerns |  |
|  | Bias in selection of the reported result | Low |  |
| 9. | Risk of bias arising from the randomization process | Some Concerns | Some Concerns |
|  | Bias due to deviations from intended intervention | Some Concerns (effect of assignment to intervention) |  |
|  |  | Low (effect of adhering to intervention) |  |
|  | Bias Due to Missing Outcome Data | Low |  |
|  | Bias in Measurement of the Outcome | Some Concerns |  |
|  | Bias in selection of the reported result | Low |  |
| 10. | Risk of bias arising from the randomization process | Some Concerns | Low Risk |
|  | Bias due to deviations from intended intervention | Low (effect of assignment to intervention) |  |
|  |  | Low (effect of adhering to intervention) |  |
|  | Bias Due to Missing Outcome Data | Low |  |
|  | Bias in Measurement of the Outcome | Some Concerns |  |
|  | Bias in selection of the reported result | Low |  |
| 16. | Risk of bias arising from the randomization process | Low | Low Risk |
|  | Bias due to deviations from intended intervention | Low (effect of assignment to intervention) |  |
|  |  | Low (effect of adhering to intervention) |  |
|  | Bias Due to Missing Outcome Data | Low |  |
|  | Bias in Measurement of the Outcome | Some Concerns |  |
|  | Bias in selection of the reported result | Low |  |

\*Note – “Some Concerns” are considered equivalent to “moderate risk”

### Synthesis and Data analysis

As outcome measures were not uniform across all studies included, a meta-analysis of the data extracted was not possible. As such, papers were compared through narrative synthesis of results, study characteristics and quality.

## RESULTS

Seventeen studies passed the screening criteria and underwent data extraction. *Table 5.* displays the characteristics of studies included. Of these, 29% assessed an already existing scheme (n=5), while 71% assessed a new therapy (n=12). The majority of studies assessed the associations of Nature-based interventions in patients with dementia (n=16), while one study enrolled patients with post-stroke fatigue. Studies originated from a range of countries; UK (n=5), Japan (n=4), USA (n=2), Netherlands, Sweden, Norway, Taiwan, China, & Australia (all n=1).

**Table 5.**
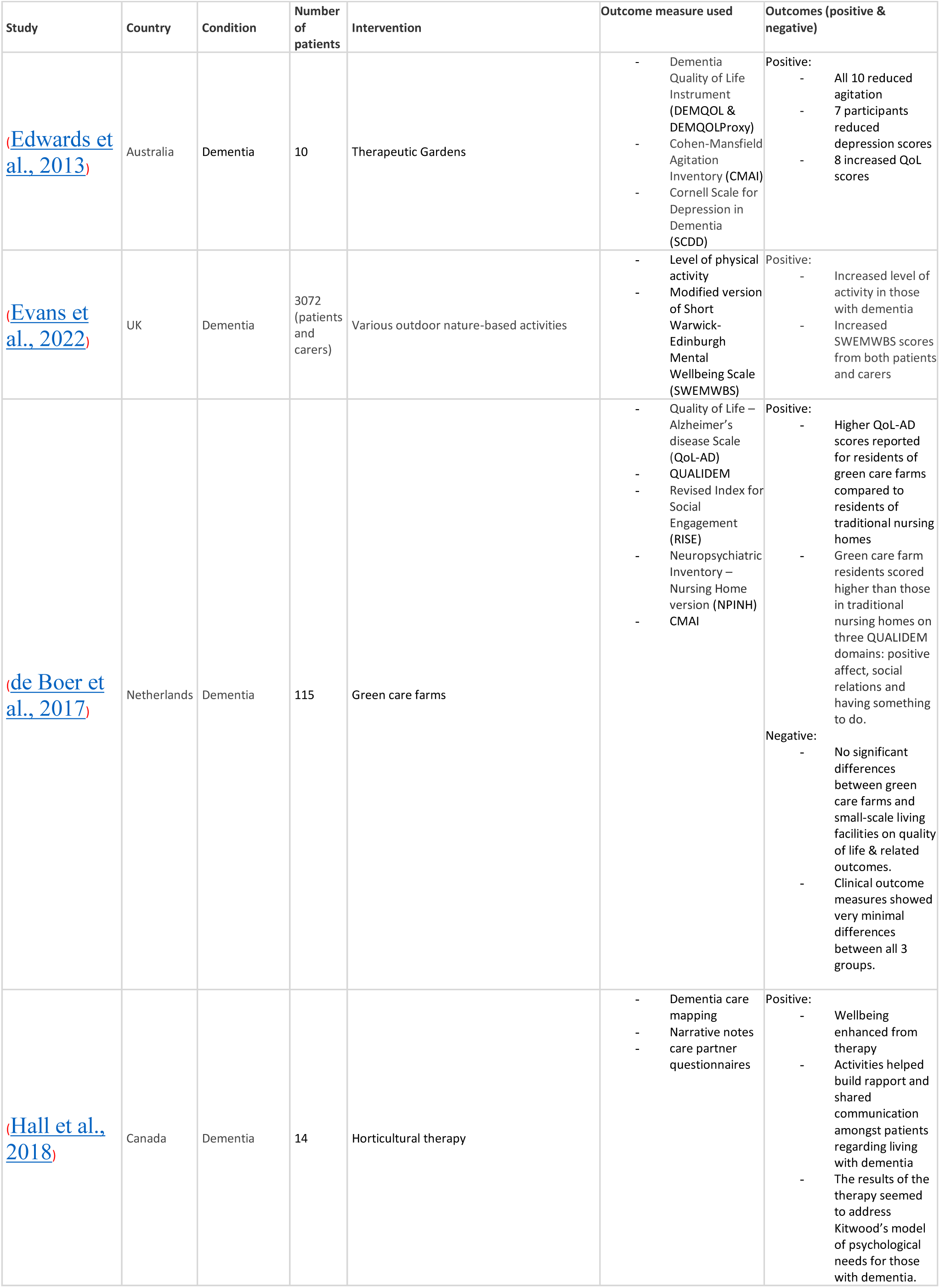
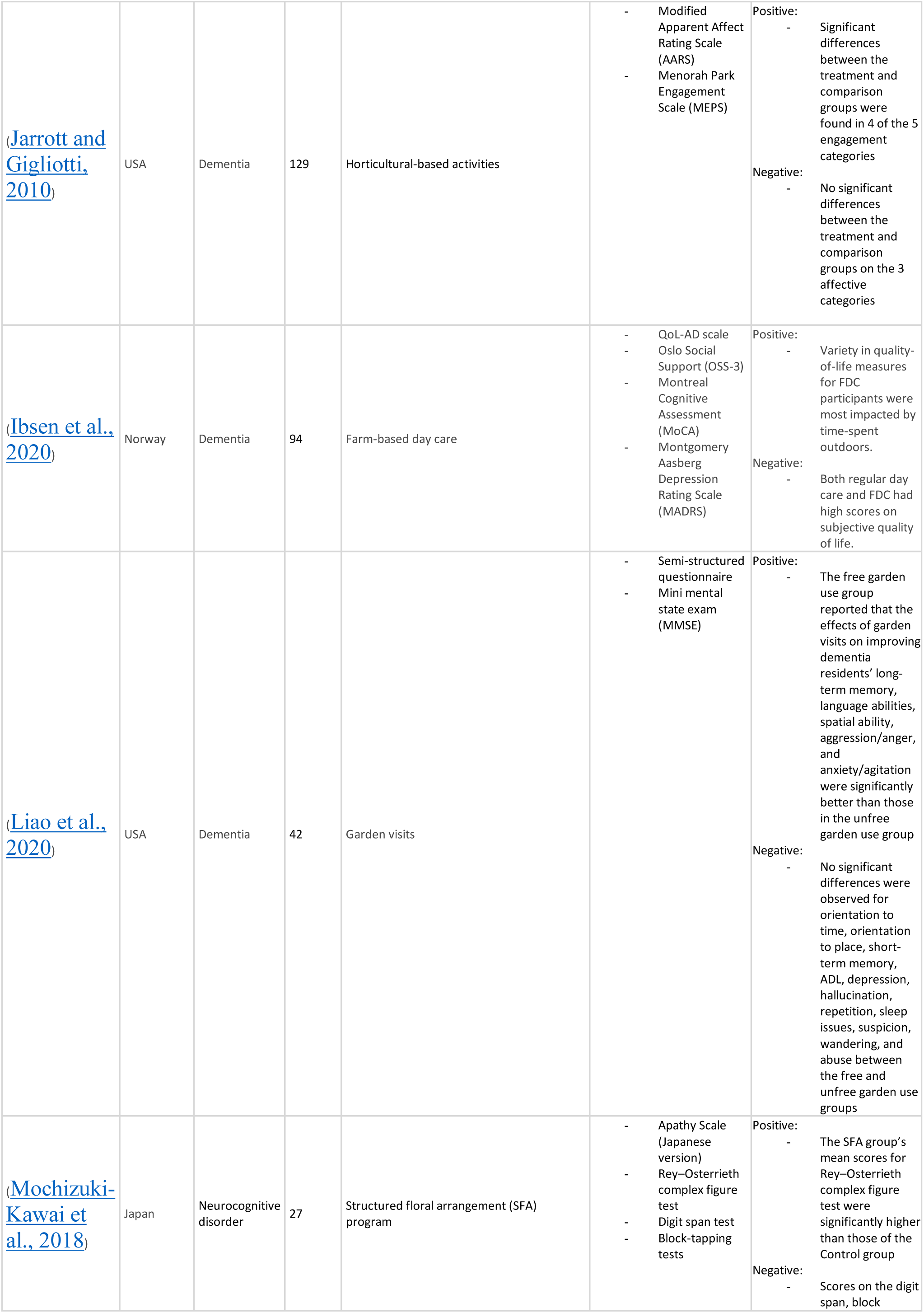
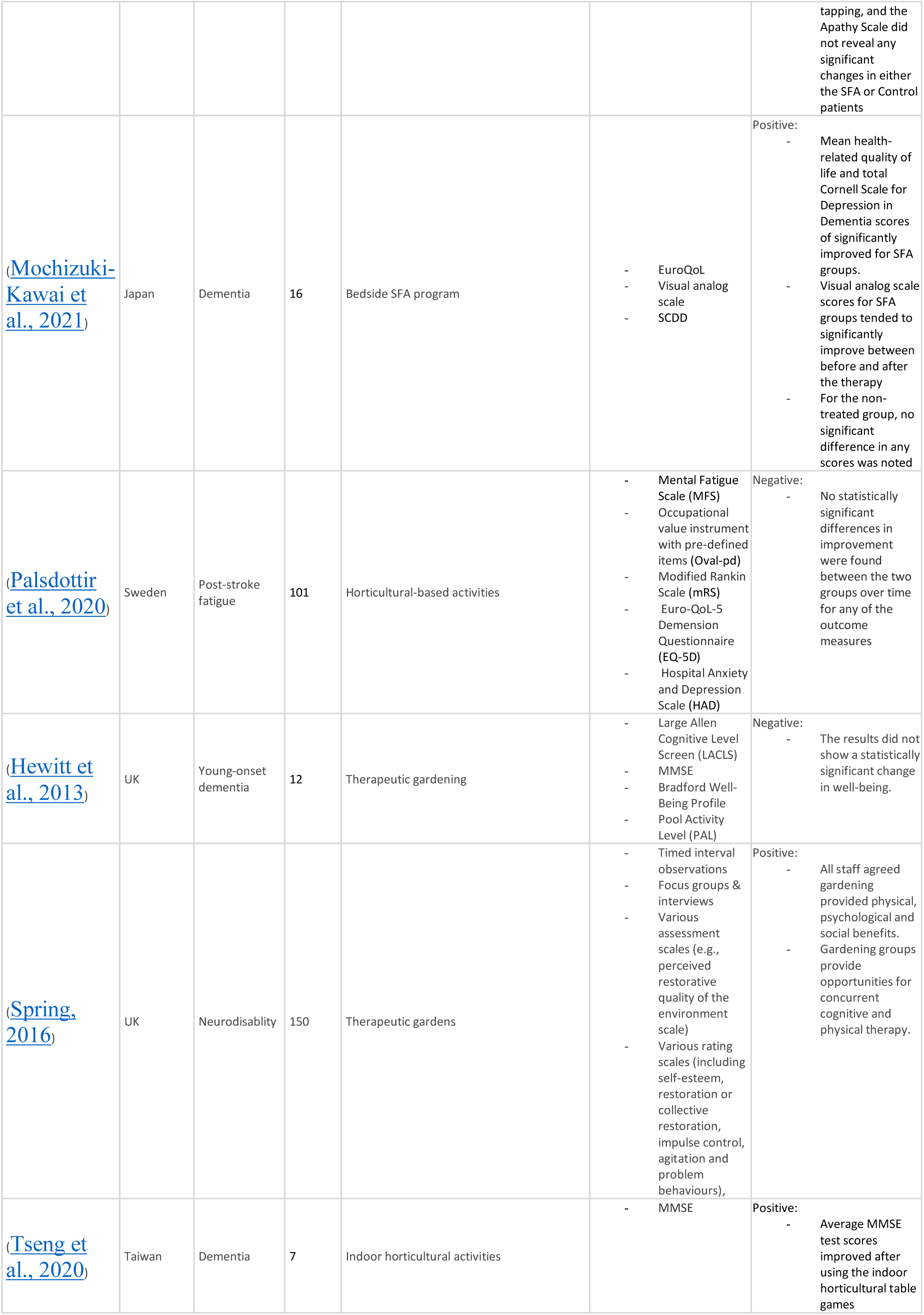
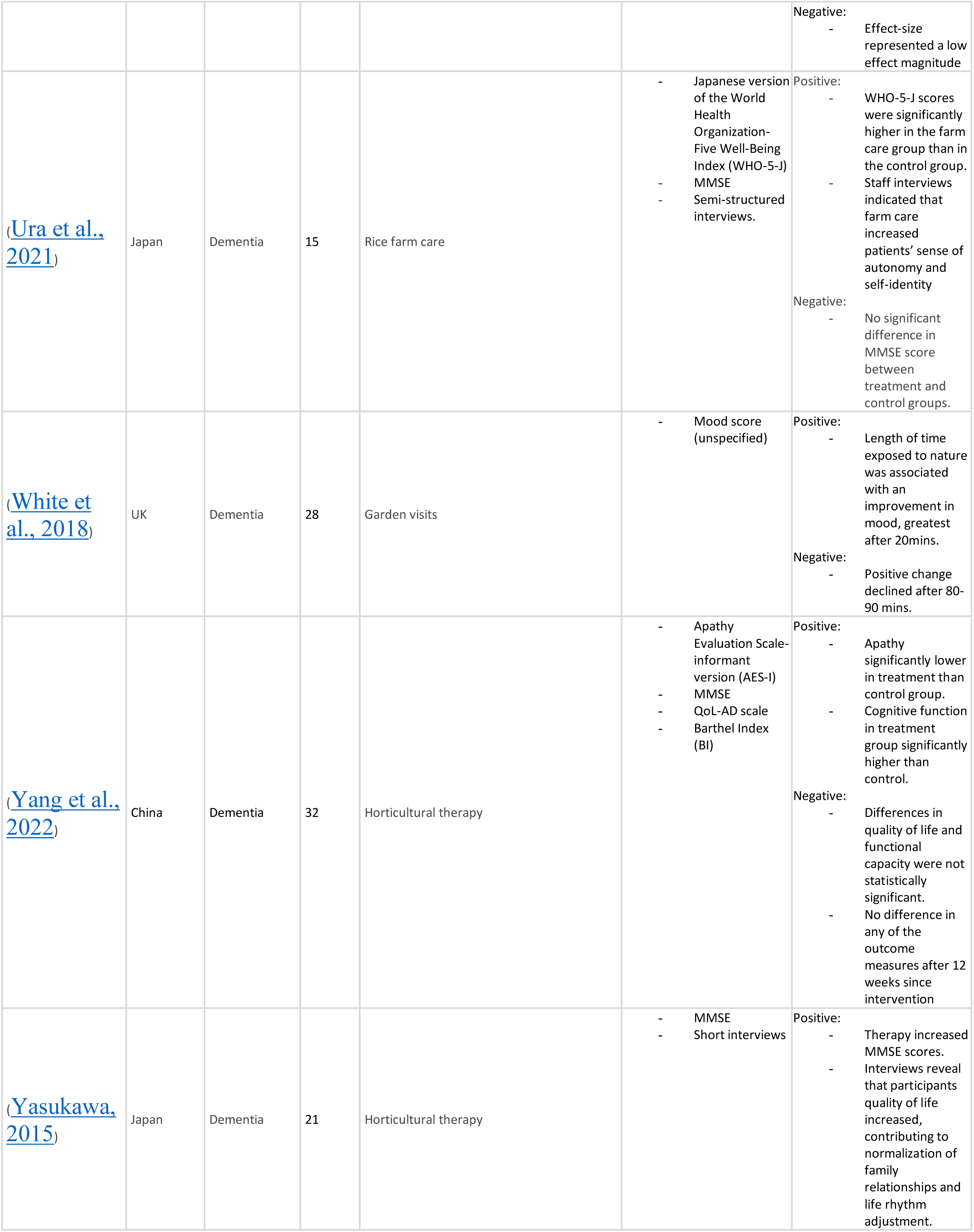
Data Extraction table.

The majority of studies (82%) were published within the last decade (n=14). Study designs consisted of quasi-experimental (n=9), randomised controlled trials (n=5) and cross-sectional (n=3). The number of participants per study varied from 7 to 3072, with the median value being 28. Only one study had a follow-up period exceeding a year. The most common intervention was horticultural therapy (n=8), while others included green care farms (n=3) and structured flower arrangement programs (n=2). Outcome measures varied between studies but most commonly incorporated quality of life assessments (n=6), interviews/questionnaires from patients and carers (n=6), or the mini-mental state exam (n=6). Other measures included wellbeing scales (n=3) and depression scores (n=3). No study used a Nature connectedness scale.

In most studies (n=15; 88%), nature-based interventions demonstrated improvements in one or more outcome measures, including quality of life, agitation, apathy, cognitive function and mood. Two studies did not report a clear beneficial outcome of stastistical significance (<u>Palsdottir et al., 2020</u>, <u>Hewitt et al., 2013</u>).

There were notable methodological inconsistencies amongst the studies. Thirteen studies included no randomisation procedure. A small sample size (<50) was found in eleven studies. Eight studies did not recruit a control/comparison group. Overall, the studies showed a wide range of bias level; three studies had high risk, eleven studies had moderate risk, and three studies had low risk. The main reason for studies being considered moderate/high-risk was the lack of a control/comparison group.

## DISCUSSION

In this systematic review, we found that 88% of studies reported at least one positive outcome when Nature-based interventions were utilised for neurological disorders, mainly in dementia. Whilst this result suggests that the integration of Nature-based interventions alongside conventional treatment approaches could improve patient outcomes and quality of life, particularly in dementia, there remain many unanswered questions. Of note, we found no studies focusing on several common neurological disorders, including migraine, epilepsy and multiple sclerosis. These conditions are highly prevalent and significantly impact the quality of life of affected individuals (<u>Gilmour et al., 2018</u>, <u>Beghi, 2020</u>). More generally, stress and psychological well-being have been identified as important factors influencing the frequency and severity of symptoms in such neurological disorders (<u>Michaelis et al., 2020</u>, <u>Pagnini et al., 2014</u>). It is noteworthy that mindfulness, which has proven its benefit in several neurological diseases, is likely to share an overlapping mechanism with many Nature-based interventions (<u>Prohaska and Matthias, 2023</u>, <u>Simpson et al., 2019</u>, <u>Lin et al., 2023</u>).

An important consideration is the heterogeneity of disease groups. Neurological disorders encompass a wide range of conditions (<u>Hussain and Shehzadi, 2023</u>), with complex neuropathology and clinical manifestations (<u>Savelieff et al., 2022</u>, <u>Li et al., 2023</u>). The effectiveness of Nature-based interventions may vary across different disorders, making it important to tailor the treatment approach to specific conditions. This heterogeneity poses a challenge in designing standardised protocols and guidelines for their use, further hindering their widespread adoption.

Limited financial resources allocated to studying these therapies has impeded robust scientific evidence to support their efficacy and safety. Despite the affordability, accessibility and the potential for a wider return on investment demonstrated by Nature-based interventions (<u>Hinde et al., 2021</u>), their underutilisation is not suprising in a healthcare industry where drug-based treatments are prioritised by stakeholders. Furthermore, bureaucratic hurdles within healthcare systems can impede the integration of Nature-based interventions into clinical practice. These hurdles involve complex regulatory processes and policy frameworks that may not currently support or incentivise the utilisation of such therapies (<u>Thompson et al., 2023</u>). Collaborative efforts between researchers and policymakers are crucial in effecting these policy changes.

Wider societal issues such as inequality, increased urban living and poverty can have an indirect effect on the accessibility and utilisation of Nature-based interventions (<u>Thompson et al., 2023</u>). Research (<u>Robinson et al., 2020</u>) has shown that deprived areas tend to have lower engagement with Nature-based activities, and individuals from these marginalised communities may face barriers accessing natural environments (<u>Hinde et al., 2021</u>), hindering their potential benefits. Consequently, the perception of healthcare professionals and the public plays a crucial role (<u>Krot and Rudawska, 2021</u>). If these therapies are not widely recognised or accepted by the medical community and the general public, they may face scepticism and resistance, hindering their integration into mainstream healthcare practices.

We argue that greater efforts should be made to improve understanding of Nature-based interventions in clinical neurology. Key approaches include training programs for healthcare professionals and awareness campaigns for the public, thereby supporting new eco-education and ecotherapy initiatives in clinical practice (<u>Lord, 2023</u>, <u>Delaney et al., 2024</u>). Furthermore, the dissemination of scientific literature and involvement of patient advocacy groups play pivotal roles in promoting the acceptance of Nature-based interventions as viable treatment options, bridging the knowledge gap and changing public perception. In order to address inequalities, we need more urban green spaces and community gardens, thereby providing more equitable access to Nature (<u>Nejade et al., 2022</u>). We hereby highlight three projects addressing these issues, namely the Cross-government Green Social Prescribing Programme (<u>NHS England</u>), the Dose of Nature charitable initiative (<u>Dose of Nature, 2020</u>), and the Royal Society for the Protection of Birds’ Nature Prescription programme (<u>RSPB Scotland, 2022</u>).

Lastly, but arguably most important of all, the challenges of climate change and environmental degradation also deserve attention. The impacts of climate change, associated with the ever-growing urbanisation of our societies, the degradation and pollution of our natural environment, and loss of biodiversity, directly affect the availability and therapeutic quality of Nature-based interventions (<u>Shanahan et al., 2019</u>). These environmental challenges highlight the urgent need to protect and restore natural environments for the well-being of both humans and the planet. Therefore, initiatives to mitigate climate change and protect natural environments should be supported.

### Strengths and Limitations

The review was strengthened by utilizing multiple databases, ensuring a more thorough and inclusive search process. The rigor of the review was enhanced through the involvement of independent reviewers and AI tools for study selection and quality assessment, minimizing potential biases in interpretation and selection.

That being said, interventions varied widely, including horticultural therapy, therapeutic gardens, and farm-based care. Many studies also used self-reported or proxy-reported measures (e.g., DEMQOL, QoL-AD, SWEMWBS), which is concerning source of potancial bias, especially given the participants being elderly &/or disabled (<u>Li et al., 2015</u>). As such, the heterogenous interventions and outcome measures used per study made comparisons across studies challenging. Other methodological limitations included a lack of control groups and small sample sizes, reducing the ability to attribute observed effects solely to the intervention. Furthermore, a majority of studies included were considered either moderate or high risk (n=14). This may impact the reliability of our findings, and is therefore imperative for future studies relating to Nature interventions to have more robust designs.

Lastly, the lack of identification of the mechanism of change in the papers across the board means that Nature as the active component is neglected in the outcome measures. For example, none of the papers measured Nature connectedness. As such, despite results echoing the presence of positive functional or organic changes from such interventions, it is difficult to discern whether these are a result of Nature itself, or due to the by-products associated with participating in Nature-based interventions (such as exercise, social connection, sense of achieving something/purpose). It could be argued that simple exposure to Nature (passive engagement) may have a positive impact, and then connecting with Nature increases this impact further. Nonetheless, in order to better investigate such relationships, future reviews and studies should focus their outcome measures to those which encourage the assessment of the extent of interacting with Nature as the main variable.

## CONCLUSION

Nature-based therapies hold great promise as complementary treatments for neurological disorders due to their cost-effectiveness, ease of accessibility, and positive health outcomes. Overcoming financial disincentives, addressing research biases, and identifying potential barriers are essential steps in advancing the integration of Nature-based interventions, including formal Nature Prescriptions, into mainstream healthcare for neurological conditions. By addressing these barriers through research, policy changes, education, and advocacy, the therapeutic potential of Nature-based therapies should be further explored. Continued collaborative efforts among various stakeholders can pave the way for a more comprehensive and patient-centred approach to managing neurological disorders, with a goal to improving patients’ overall well-being and quality of life.

## Data Availability

All data produced in the present study are available upon reasonable request to the authors.
All data produced in the present work are contained in the manuscript.

---

**APPENDIX.** A. Full search Strategy.

A. Full search Strategy
| PEO elements | Key words | Search terms | Search strategy |
| --- | --- | --- | --- |
| **Population** | Patients with common neurological disease | stroke, migraine, epilepsy, multiple sclerosis, neurodegenerative disease, myasthenia gravis, brain tumour, brain injury, headache, functional neurological disorder. | Neurological disease OR Neurologic disease OR Stroke OR Migraine OR Epilepsy OR Multiple sclerosis OR MS OR neurodegenerative disease OR Parkinson's disease OR PD OR Alzheimer's disease OR AD OR Dementia OR ALS OR Amyotrophic Lateral Sclerosis OR MND OR Motor neurone disease OR Lou Gehrig's disease OR Functional Neurological Disorder OR FND OR Seizure OR Headache OR Myasthenia gravis OR MG<br>OR Brain injury OR Brain tumour OR Neuropathy |
| **Exposure/intervention** | Nature-based treatments | ecotherapy, wilderness therapy, adventure therapy, green prescriptions, nature-based interactions. | AND<br>ecotherapy OR nature therapy OR ecological therapy OR wilderness therapy OR adventure therapy OR green prescriptions OR green prescribing OR nature-based interactions OR nature-based treatments OR nature exposure OR natural environment OR green space OR Horticultural therapy OR Green exercise OR Nature-based interventions OR Nature-associated interventions OR Animal assisted therapy OR Care farming OR Social farming OR Green care |
| **Outcome measure** | Improved health outcomes | cognitive function, brain activity, blood pressure, mental health, physical activity, sleep | AND<br>Health outcomes OR cognitive function OR brain activity OR blood pressure OR mental health OR physical activity OR sleep OR life impact OR quality of life OR QOL OR Functional rating OR Well-being OR wellbeing OR Physical health OR disease progression OR health status |

